# Elucidating the genetic risk of obesity through the human blood plasma proteome

**DOI:** 10.1101/2020.05.31.20118208

**Authors:** Shaza B. Zaghlool, Sapna Sharma, Megan Molnar, Pamela R. Matías‐García, Mohamed A. Elhadad, Melanie Waldenberger, Annette Peters, Wolfgang Rathmann, Johannes Graumann, Christian Gieger, Harald Grallert, Karsten Suhre

**Affiliations:** Department of Physiology and Biophysics, Weill Cornell Medicine‐Qatar, Doha, Qatar.; Research Unit of Molecular Epidemiology, Helmholtz Zentrum München, German Research Center for Environmental Health, Neuherberg, Bavaria, Germany.; Institute of Epidemiology, Helmholtz Zentrum München, German Research Center for Environmental Health, Neuherberg, Bavaria, Germany.; German Center for Diabetes Research (DZD), Ingolstädter Landstr. 1, 85764 Neuherberg, Germany; TUM School of Medicine, Technical University of Munich, Munich, Germany; German Centre for Cardiovascular Research (DZHK), partner site Munich Heart Alliance, Munich, Germany.; German Research Center for Cardiovascular Research (DZHK), partner site Munich Heart Alliance, Germany.; Institute of Biometrics and Epidemiology, German Diabetes Center, Düsseldorf, Germany.; Scientific Service Group Biomolecular Mass Spectrometry, Max Planck Institute for Heart and Lung Research, W.G. Kerckhoff Institute, Bad Nauheim, Germany.; German Centre for Cardiovascular Research (DZHK), partner site Rhine‐Main, Max Planck Institute of Heart and Lung Research, Bad Nauheim, Germany.

**Keywords:** blood proteome, association study, polygenic score, body mass index, obesity, Mendelian randomization

## Abstract

Obesity is affecting an increasing number of individuals worldwide, but the complex interplay between genetic, environmental and lifestyle factors that control body weight is still poorly understood. Blood circulating protein are confounded readouts of the biological processes that occur in different tissues and organs of the human body. Many proteins have been linked to complex disorders and are also under substantial genetic control. Here, we investigate the associations between over 1,000 blood circulating proteins and body mass index (BMI) in three studies, including over 4,600 participants. We show that BMI is associated with widespread changes in the plasma proteome. We report 152 protein associations with BMI that replicate in at least one other study. 24 proteins also associate with a genome‐wide polygenic score (GPS) for BMI. These proteins are involved in lipid metabolism and inflammatory pathways impacting clinically relevant pathways of adiposity. Mendelian randomization suggests a bi‐directional causal relationship of BMI with three proteins (LEPR, IGFBP1, and WFIKKN2), a protein‐to‐BMI relationship for three proteins (AGER, DPT, and CTSA), and a BMI‐to‐protein relationship for 21 other proteins. Combined with animal model and tissue‐specific gene expression data, our findings suggest potential therapeutic targets and further elucidate the biological role of these proteins in pathologies associated with obesity.

## INTRODUCTION

Obesity is a multifactorial disorder with poorly understood causative mechanisms and a large polygenic contribution [1]. Genome‐wide association studies of BMI identified genetic variants that can account for ∼2.7% ‐ 6% of observed variance in BMI [2, 3]. Due to an increasingly sedentary lifestyle and a food industry transition to processed food, the prevalence of obesity worldwide has tripled over the past 40 years [4]. Based on the latest estimates in European Union countries, 30‐70% of adults are overweight and 10‐30% are obese (World Health Organization). Obesity greatly increases the risk of several chronic diseases, such as depression, type 2 diabetes, cardiovascular disease, and certain cancers, putting a great burden on the healthcare system. Therefore, a better understanding of the interaction between lifestyle choices, environmental factors, and genetic predispositions is critical for the development of effective treatments and preventive interventions [5, 6].

Genetic composition is determined at conception and can be used to make predictions regarding disease susceptibility. The dramatic increase in obesity rates clearly points toward non‐genetic factors or environmental factors as major drivers, most likely in interaction with genetic variants [7]. Although some diseases can result from a single rare monogenic mutation with a large effect, most common diseases are the consequence of a cumulative effect of polygenic inheritance encompassing numerous variants, each making only a small contribution [8]. Through genome-wide association studies (GWAS), more than 900 genetic variants have been identified to be associated with obesity [3]. Although GWAS have effectively mapped these associations, the explained variance still does not fully explain the molecular mechanisms behind obesity. Genome-wide polygenic scores (GPS) are currently being used to quantify inherited disease susceptibility [9] and can explain ∼13.9% of the variance in BMI, which is more than twice the variance in BMI that can be explained using only the GWAS loci [3]. These scores approximate a normal distribution in the population and there is considerable correlation between GPS and BMI. Individuals in the upper tail of the GPS distribution can be susceptible to genetic effects comparable to carriers of single rare monogenic disease variants [9].

Given that proteins are the main building blocks of an organism and potential drug targets, proteome‐wide association analysis seems to be the obvious next step in obesity research [10]. Levels of many proteins vary significantly between obese and normal individuals [11, 12]. Until recently, mass‐spectrometry‐based proteomic analyses of blood samples were limited to small sample sizes or a limited number of measured proteins. Multiplexed affinity‐based proteomics approaches using antibodies or specifically designed aptamers now allow quantification of levels of hundreds of proteins from small amounts of plasma or serum sample. We previously quantified 1,100 blood circulating proteins using the SOMAscan affinity proteomics platform (Somalogic Inc.) [13] in samples from 996 individuals of the population‐based KORA F4 (Cooperative Health Research in the Region of Augsburg) study [14].

Here, we report a high throughput proteomics association study with BMI in KORA (Germany), and a replication in two independent studies, including 356 participants of the multi‐ethnic Qatar Metabolomics Study on Diabetes (QMDiab) [15], and publicly available association statistics from 3,301 individuals of the INTERVAL study (UK) [16]. We compute genome‐wide polygenic scores (GPS) for BMI [17] and identify proteins whose levels associate with the GPS for BMI. We then use Mendelian randomization paired with experimental evidence to identify proteins and pathways that may be causally linked to obesity. The study design and summary results are presented in Figure 1.

**Figure 1:**
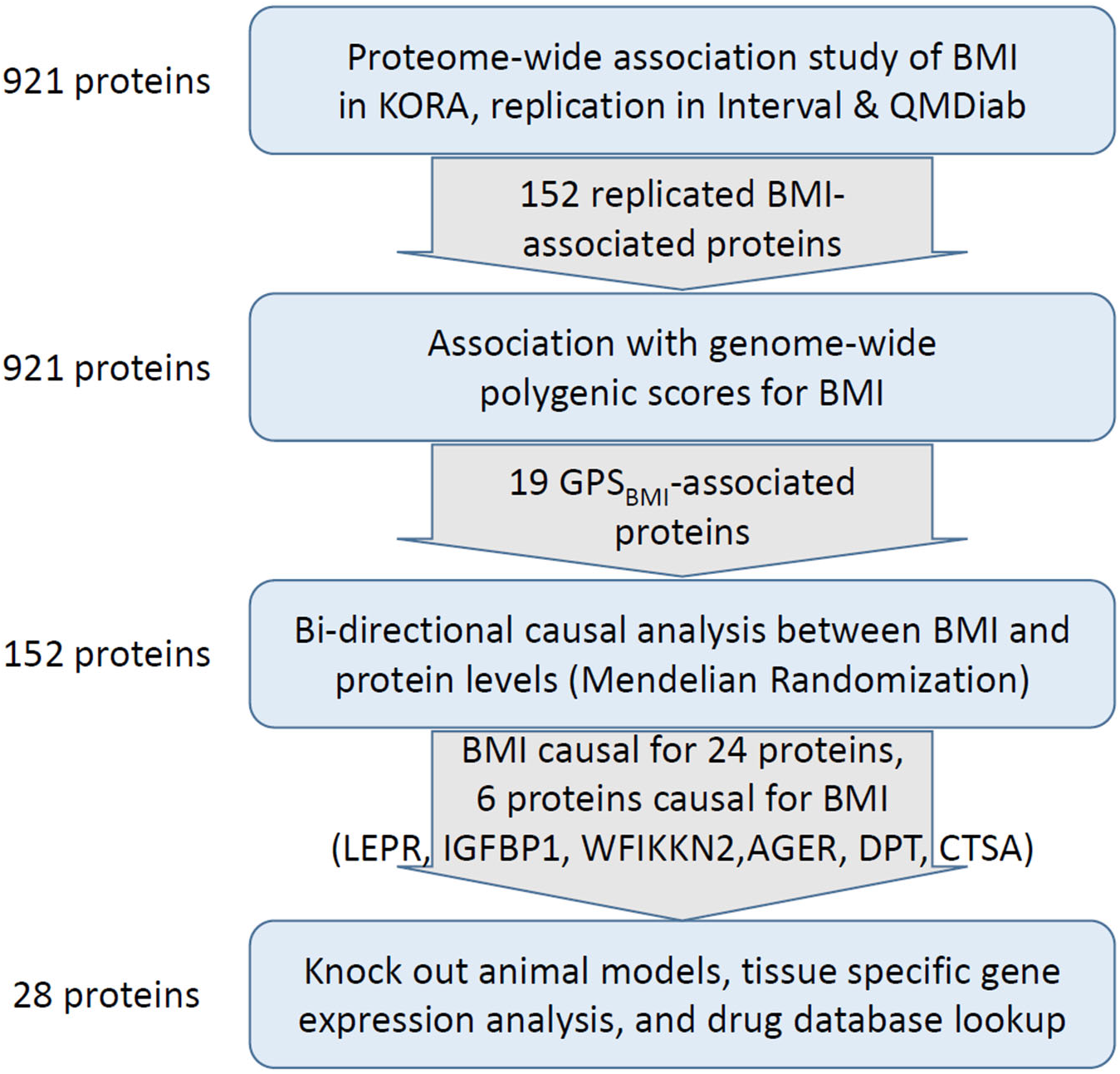
Study Overview. (top) Protein-wide association study with BMI conducted in KORA with confirmation/replication in INTERVAL and QMDiab; (top-middle) Protein-wide association study with BMI polygenic scores (in KORA and QMDiab); (bottom-middle) Bi-directional causality analysis to determine if BMI has a potentially causal effect on protein levels and/or if proteins are potentially causal in the development of obesity; (bottom) Studying tissue-specific gene expression in humans/mice, identification of genes encoding proteins related to obesity traits, searching for existing animal models, and identification of potential targetable proteins through drug database searches.

## RESULTS

### 152 out of 921 assayed blood protein levels associate with body mass index

After stringent quality control, we identified 921 proteins whose levels were determined in 996 blood samples from the KORA study and that were also measured, both in the INTERVAL and the QMDiab studies. Although not available for all three studies, we also included the leptin (LEP) and leptin receptor (LEPR) proteins for their well‐studied roles in obesity. The study descriptive statistics for the 996 individuals are provided in Supplementary Table 1. We used linear regression including age and sex as covariates to carry out a protein‐wide association in KORA and identified 184 associations between log2 transformed blood circulating protein levels and BMI after conservative Bonferroni correction (p < 5.43×10^‐5^; 0.05/921). 107 proteins were negatively correlated with BMI while 77 were positively correlated (Figure 2 and Supplementary Table 2).

**Figure 2:**
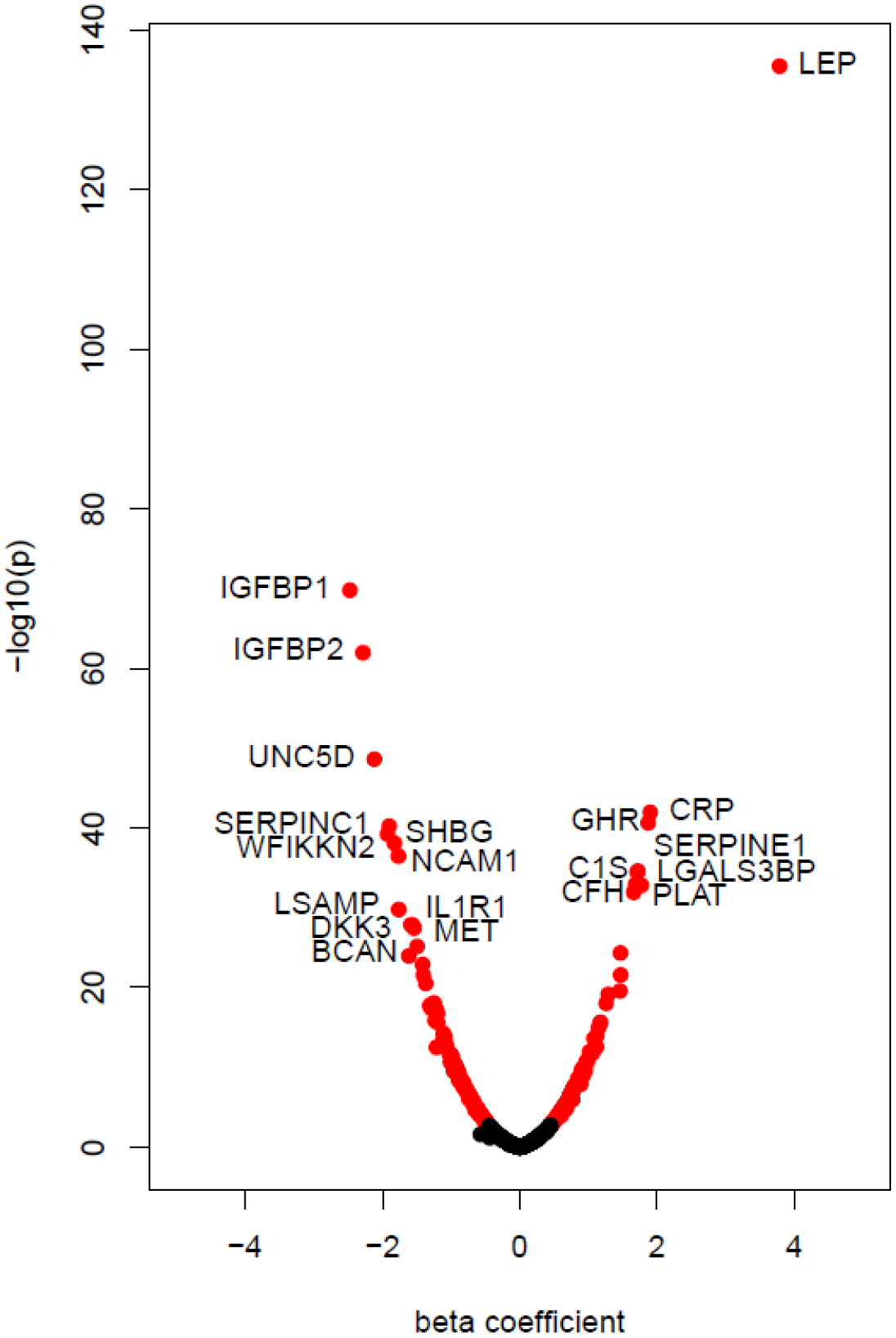
Protein-wide association study with body mass index. Volcano plot showing the association of BMI with plasma protein levels in KORA, including age and sex as covariates. Leptin is the strongest protein associated with BMI (p = 3.34×10^−136^) in addition to 151 significantly associated proteins (red).

We tested the influence of a number of potential confounders on the BMI‐protein associations, specifically, smoking status, alcohol consumption, physical activity, and binary diabetes state (Supplementary Table 3). We did not observe any substantial effect of confounding by these factors on the BMI‐protein association results, and all Bonferroni significant protein‐BMI associations found in the full model were also significant in the model where only age and sex were included as covariates.

We confirmed the BMI‐protein associations using the published INTERVAL associations [16] and additionally attempted replication of these 184 associations in the multi‐ethnic QMDiab study (Supplementary Table 4). Of the 184 proteins, 150 BMI‐protein associations (81.5%) were significant in INTERVAL, after Bonferroni correction (p < 2.72×10^‐4^; 0.05 / 184). In QMDiab, 37 (20.1%) of the BMI‐protein associations were replicated after Bonferroni correction, while a further 131 proteins (71.2%) were directionally concordant, but were not sufficiently powered for replication. In total, 152 BMI‐protein associations were replicated in at least one study – specifically, 35 associations replicated in both studies, 115 associations replicated only in INTERVAL, and two new associations replicated only in QMDiab (THBS2 and ANGPT2). In addition, we found that out of 28 BMI‐protein associations that had 95% replication power (determined by sampling), 17 proteins (60.7%) fully replicated in QMDiab, and 26 proteins (92.9%) displayed at least nominal significance. The Pearson correlation for the effect sizes is R = 0.92 between KORA and INTERVAL, and R = 0.84 between KORA and QMDiab. (Supplementary Figure 1).

### Association of BMI polygenic scores with BMI

We computed genome‐wide polygenic scores for BMI (GPS_BMI_) for 996 participants of KORA and 353 of QMDiab using variants and weights as described previously [9, 17]. Briefly, the GPS_BMI_ score is based on summary statistics from a recent genome‐wide association study (GWAS) with BMI and assigns weights to each genetic variant depending on the strength of its association with BMI (see Methods). The GPS_BMI_ was strongly associated with BMI in KORA (p = 2.32 × 10^‐43^), and was also significant in the multi‐ethnic QMDiab study (p = 5.54 × 10^‐4^) (Supplementary Figures 2a and 2b).

19 proteins were associated with GPS_BMI_ in KORA after accounting for multiple testing (p < 5.43 × 10^‐5^; 0.05/921) (Supplementary Table 5). All 19 proteins were also strongly associated with BMI in KORA (Table 1). The strongest protein association with BMI (LEP; p = 3.34×10^‐136^) was also the strongest with GPS_BMI_ (p = 1.32×10^‐12^).

**Table 1:**
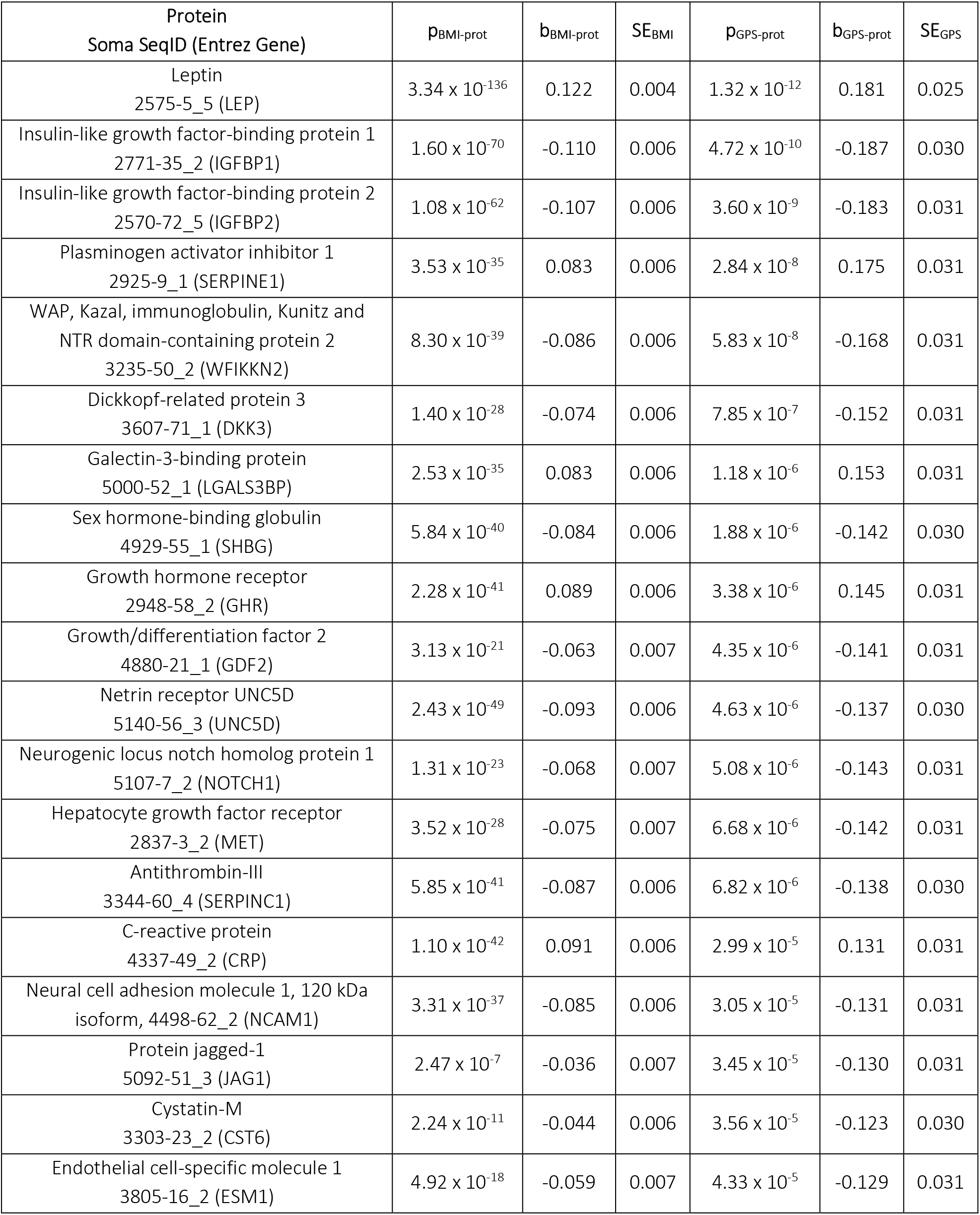
Proteins significantly associated with BMI and the polygenic score for BMI. The p-values (p_BMI-prot_), regression coefficients (b_BMI-prot_) are for the BMI-protein associations, while p_GPS-prot_ and bGPS-prot are for the GPS_BMI_-protein associations.

We replicated the analysis in QMDiab to evaluate the applicability of a polygenic score derived from European participants to a cohort of mixed non‐Caucasian ethnicity. Using linear regression and adjusting for age, sex, and study‐specific covariates (described in Methods), five log 2 transformed proteins remained significantly associated with GPS_BMI_ after Bonferroni correction in QMDiab (p<0.05/19; 2.63×10^‐3^) (NOTCH1, C5, NCAM1, CRP, and SERPINC1), while another six proteins (LEP, IGFBP1, WFIKKN2, UNC5D, MET, RARRES2), were nominally associated with concordant directionality (p<0.05) (Supplementary Table 6).

To confirm that the GPS_BMI_ to protein associations were truly polygenic, as opposed to potentially being driven by a few strong *in‐cis* variant effects, we excluded all genetic variants within 1 MB of the genes encoding the associated protein from score computation. All of the 19 protein associations with GPS_BMI_ remained significant after eliminating potential cis‐pQTL effects (Supplementary Table 7).

### Extreme BMI polygenic scores identify 19 proteomic signatures for 5% of the population

Interestingly, the association between GPS_BMI_ and BMI was not linear. The effect estimate was in fact much stronger at the extremes of the distribution (Figure 3) and agrees with previous reports [9]. To evaluate this “tail‐effect” in our study, we stratified the 996 KORA study samples based on GPS_BMI_ percentiles. We found a steeper slope with respect to both BMI and leptin (LEP) measures at the lower and upper extremes of the distribution. For instance, the mean BMI was 24.94 kg/m^2^ [CI = 24.29; 25.60] in the bottom decile and 31.09 kg/m^2^ [CI = 31.09; 32.21] in the top decile translating to a significant difference between the groups (two sample t‐test p = 3.82×10^‐17^).

**Figure 3:**
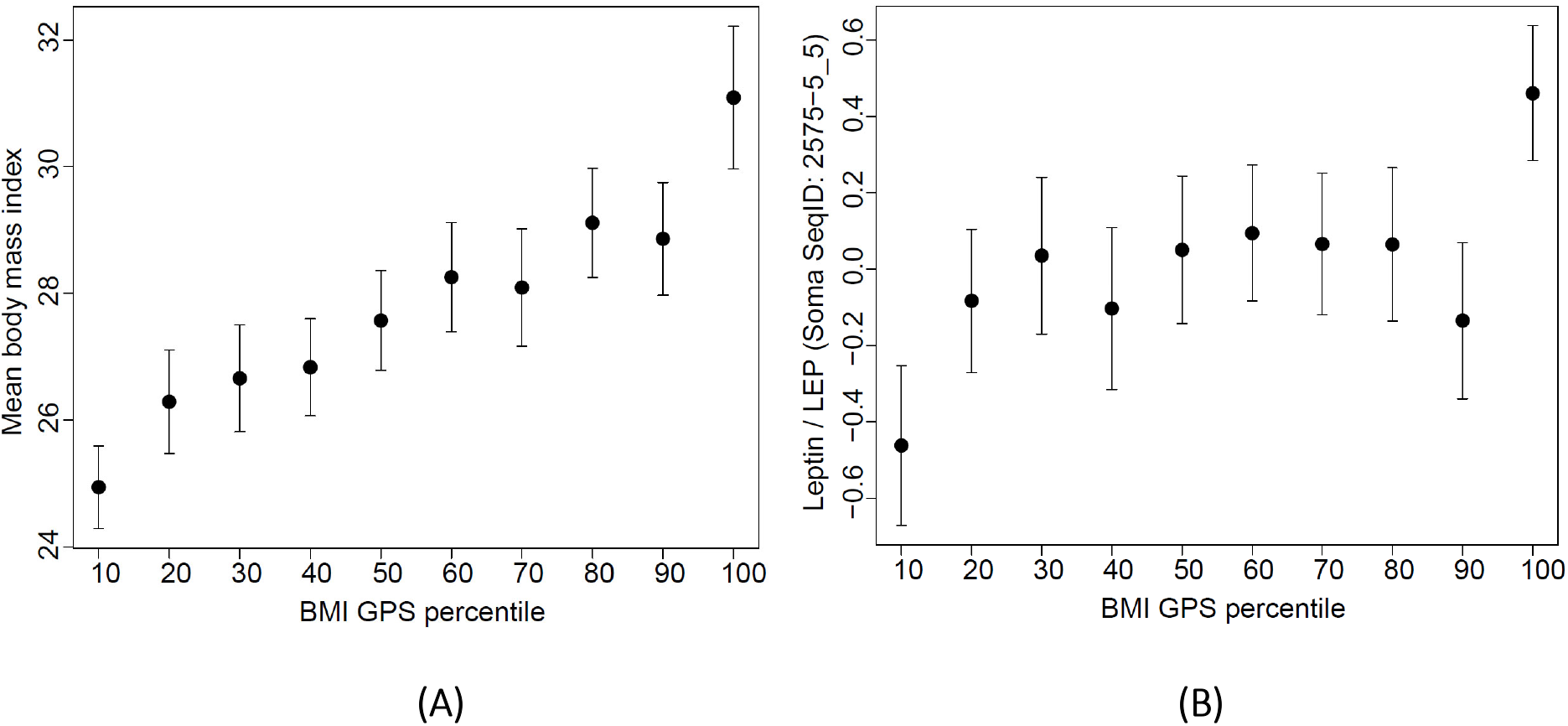
Stratification of the KORA samples according to GPS_BMI_ percentiles. There is a particularly steep slope with respect to both, (A) BMI and (B) leptin (LEP) measures at the upper and lower deciles.

To investigate whether a similar “tail‐effect” can be observed for the associations between GPS_BMI_ and blood circulating proteins, we compared the different effect sizes and significance levels at various percentiles of the GPS_BMI_ distribution (Supplementary Table 8), including the full dataset (N = 996), the 25^th^ vs. 75^th^ percentiles, the 20^th^ vs. 80^th^ percentiles, the 15^th^ vs. 85^th^ percentiles, the 10^th^ vs. 90^th^ percentiles, and the 5^th^ vs. 95^th^ percentiles. We found that the effect of GPS_BMI_ on the log2 transformed leptin was almost quadrupled in the 5% tail of the population compared to the full data (Figure 4). Individuals in the extreme tail of the GPS_BMI_ distribution showed an over-proportionally increased genetic predisposition for developing obesity [17]. We found a similar effect for the 19 GPS_BMI_‐associated proteins (Table 2) (Supplementary Figure 3).

**Figure 4:**
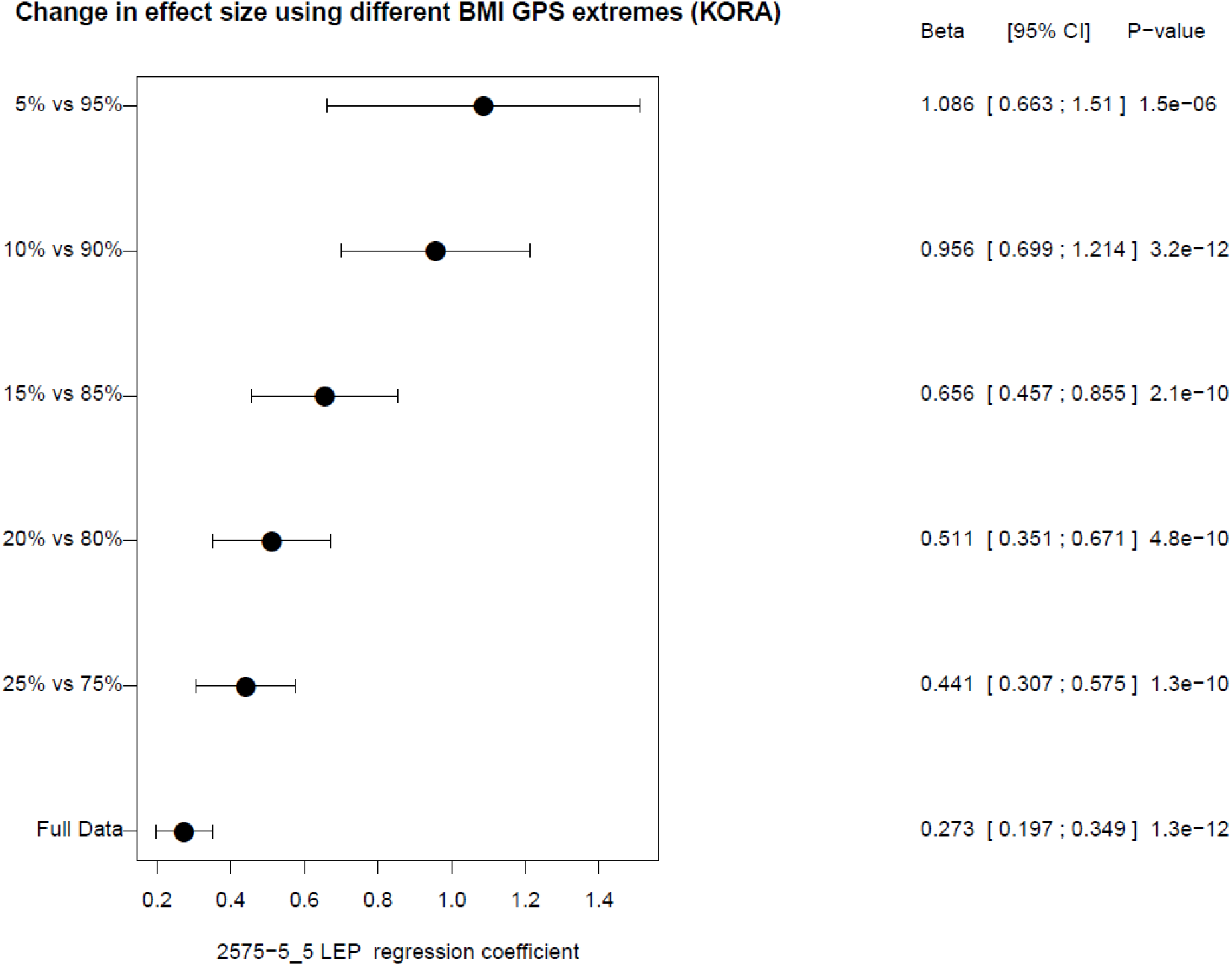
Extreme GPS_BMI_ is a strong risk factor for increased leptin levels and severe obesity. The effect of GPS_BMI_ on leptin is almost quadrupled in the extreme 5% of the population compared to the full data (N = 996).

**Table 2:** The over-proportional contribution of genetics to BMI in the tail of the GPSBMI distribution translates to at least a 3-fold increase/decrease in protein levels. The effect sizes (beta) and p-values (P) are presented for the full data set and limited to data in the 5% tails of the GPSBMI, respectively.

| Protein | Beta<sub>full</sub> | P<sub>full</sub> | Beta<sub>5%</sub> | P<sub>5%</sub> | $\frac{Beta_{5\%}}{Beta_{full}}$ |
| --- | --- | --- | --- | --- | --- |
| IGFBP2 | -0.189 | 3.60 x 10<sup>-9</sup> | -1.073 | 1.08 x 10<sup>-7</sup> | 5.671 |
| IGFBP1 | -0.205 | 4.72 x 10<sup>-10</sup> | -1.046 | 1.49 x 10<sup>-6</sup> | 5.101 |
| LEP | 0.273 | 1.32 x 10<sup>-12</sup> | 1.086 | 1.52 x 10<sup>-6</sup> | 3.977 |
| SERPINE1 | 0.175 | 2.84 x 10<sup>-8</sup> | 1.002 | 4.07 x 10<sup>-6</sup> | 5.732 |
| UNC5D | -0.152 | 4.63 x 10<sup>-6</sup> | -1.102 | 6.89 x 10<sup>-6</sup> | 7.230 |
| WFIKKN2 | -0.174 | 5.83 x 10<sup>-8</sup> | -0.908 | 7.83 x 10<sup>-6</sup> | 5.227 |
| SHBG | -0.159 | 1.88 x 10<sup>-6</sup> | -0.958 | 1.43 x 10<sup>-5</sup> | 6.014 |
| NCAM1 | -0.132 | 3.05 x 10<sup>-5</sup> | -0.723 | 1.85 x 10<sup>-4</sup> | 5.464 |
| NOTCH1 | -0.145 | 5.08 x 10<sup>-6</sup> | -0.728 | 2.16 x 10<sup>-4</sup> | 5.011 |
| MET | -0.143 | 6.68 x 10<sup>-6</sup> | -0.718 | 2.27 x 10<sup>-4</sup> | 5.034 |
| DKK3 | -0.160 | 7.85 x 10<sup>-7</sup> | -0.857 | 2.67 x 10<sup>-4</sup> | 5.360 |
| SERPINC1 | -0.147 | 6.82 x 10<sup>-6</sup> | -0.793 | 7.65 x 10<sup>-4</sup> | 5.408 |
| CST6 | -0.139 | 3.56 x 10<sup>-5</sup> | -0.818 | 8.66 x 10<sup>-4</sup> | 5.894 |
| JAG1 | -0.132 | 3.45 x 10<sup>-5</sup> | -0.701 | 1.35 x 10<sup>-3</sup> | 5.294 |
| LGALS3BP | 0.154 | 1.18 x 10<sup>-6</sup> | 0.715 | 1.69 x 10<sup>-3</sup> | 4.651 |
| GDF2 | -0.149 | 4.35 x 10<sup>-6</sup> | -0.676 | 4.40 x 10<sup>-3</sup> | 4.524 |
| GHR | 0.148 | 3.38 x 10<sup>-6</sup> | 0.570 | 7.62 x 10<sup>-3</sup> | 3.841 |
| ESM1 | -0.130 | 4.33 x 10<sup>-5</sup> | -0.514 | 2.00 x 10<sup>-2</sup> | 3.952 |
| CRP | 0.133 | 2.99 x 10<sup>-5</sup> | 0.406 | 5.11 x 10<sup>-2</sup> | 3.044 |

### Mendelian Randomization

To assess whether proteins are causally affected by BMI in the direction (BMI‐to‐protein) or vice versa (protein‐to‐BMI), we carried out bi‐directional Mendelian randomization investigations. We initially conducted both, a one‐sample (1SMR) and a two‐sample (2SMR) Mendelian randomization analysis, and in both directions (Table 3). In the BMI‐to‐protein direction, we used GPS_BMI_ as an instrument for BMI. Our results indicated that the 1SMR had higher statistical power than the 2SMR in identifying significant MR associations. This is plausible, because the BMI instrument was generated using variant weights from the largest GWAS with BMI and sample level data and the 1SMR used individual‐level data, while the 2SMR only had access to protein summary statistics from a study that is merely four times the size of KORA.

**Table 3:** Accumulative evidence is suggestive of relationships between BMI and proteins in both
directions. MR analysis is summarized for both directions (BMI-to-protein and protein-to-BMI) for one-sample MR (1SMR) using the 2SLS method, and two-sample MR (2SMR) using the IVW method. Entries in red are Bonferroni significant (corrected for the respective number of MR tests).

| Direction | BMI→Protein |  |  |  | Protein→BMI |  |  |  | Animal model |
| --- | --- | --- | --- | --- | --- | --- | --- | --- | --- |
| Method | 1SMR (KORA) |  | 2SMR (GIANT+UKBiobank / INTERVAL) |  | 1SMR (KORA) |  | 2SMR (PheWeb browser) |  |  |
| Protein | Beta | P | Beta | P | Beta | P | Beta | P |  |
| LEP<br>2575-5_5 | 0.104 | 1.43x10<sup>-8</sup> | 0.424 | 1.01x10<sup>-6</sup> | No instruments |  | No instruments |  | yes |
| LEPR<br>5400-52_3 | Not measured in KORA |  | -0.212 | 1.14x10<sup>-2</sup> | Not measured in KORA |  | 0.007 | 4.62x10<sup>-3</sup> | yes |
| IGFBP1<br>2771-35_2 | -0.123 | 7.47x10<sup>-12</sup> | -0.225 | 8.05x10<sup>-3</sup> | No instruments |  | -0.053 | 5.51x10<sup>-4</sup> | yes |
| IGFBP2<br>2570-72_5 | -0.121 | 9.73x10<sup>-11</sup> | -0.277 | 9.48x10<sup>-4</sup> | No instruments |  | No instruments |  | yes |
| SERPINE1<br>2925-9_1 | 0.109 | 3.04x10<sup>-8</sup> | 0.208 | 1.62x10<sup>-2</sup> | No instruments |  | No instruments |  | no |
| WFIKKN2<br>3235-50_2 | -0.109 | 2.49x10<sup>-8</sup> | -0.290 | 5.00x10<sup>-4</sup> | -0.134 | 0.749 | -0.016 | 7.39x10<sup>-5</sup> | yes |
| DKK3<br>3607-71_1 | -0.102 | 4.70x10<sup>-7</sup> | -0.235 | 5.19x10<sup>-3</sup> | -1.226 | 0.064 | -0.009 | 0.246 | no |
| LGALS3BP<br>5000-52_1 | 0.092 | 2.24x10<sup>-6</sup> | Data not available |  | No instruments |  | 0.018 | 0.279 | yes |
| SHBG<br>4929-55_1 | -0.091 | 1.85x10<sup>-6</sup> | Data not available |  | No instruments |  | 0.014 | 0.233 | yes |
| GHR<br>2948-58_2 | 0.095 | 1.18x10<sup>-6</sup> | 0.212 | 1.11x10<sup>-2</sup> | No instruments |  | No instruments |  | yes |
| GDF2<br>4880-21_1 | -0.094 | 4.12x10<sup>-6</sup> | Data not available |  | No instruments |  | No instruments |  | yes |
| UNC5D<br>5140-56_3 | -0.092 | 1.08x10<sup>-6</sup> | -0.197 | 2.25x10<sup>-2</sup> | No instruments |  | -0.001 | 0.978 | no |
| NOTCH1<br>5107-7_2 | -0.091 | 4.98x10<sup>-6</sup> | -0.233 | 5.44x10<sup>-3</sup> | -0.257 | 0.758 | No instruments |  | yes |
| MET<br>2837-3_2 | -0.085 | 1.36x10<sup>-5</sup> | -0.087 | 0.296 | 0.002 | 0.997 | No instruments |  | yes |
| SERPINC1<br>3344-60_4 | -0.081 | 2.60x10<sup>-5</sup> | -0.057 | 0.491 | No instruments |  | No instruments |  | no |
| CRP<br>4337-49_2 | 0.074 | 1.25x10<sup>-4</sup> | 0.314 | 2.40x10<sup>-4</sup> | No instruments |  | -0.012 | 0.205 | no |
| NCAM1<br>4498-62_2 | -0.082 | $2.43 \times 10^{-5}$ | -0.304 | $3.25 \times 10^{-4}$ | No instruments | | 0.015 | 0.197 | no |
| JAG1<br>5092-51_3 | -0.086 | $5.40 \times 10^{-5}$ | -0.195 | $2.22 \times 10^{-2}$ | -0.312 | 0.818 | -0.011 | 0.321 | no |
| CST6<br>3303-23_2 | -0.071 | $6.56 \times 10^{-4}$ | -0.084 | 0.320 | No instruments | | No instruments | | no |
| ESM1<br>3805-16_2 | -0.086 | $3.27 \times 10^{-5}$ | -0.255 | $2.33 \times 10^{-3}$ | No instruments | | No instruments | | no |
| ADIPOQ<br>3554-24_1 | -0.078 | $9.14 \times 10^{-5}$ | Data not available | | No instruments | | No instruments | | yes |
| IBSP<br>3415-61_2 | -0.079 | $1.13 \times 10^{-4}$ | No instruments | | No instruments | | No instruments | | no |
| C1S<br>3590-8_3 | 0.086 | $9.67 \times 10^{-6}$ | Data not available | | 0.869 | 0.028 | No instruments | | no |
| POSTN<br>3457-57_1 | -0.081 | $1.87 \times 10^{-4}$ | No instruments | | -1.519 | 0.077 | No instruments | | yes |
| IGHM<br>3069-52_3 | -0.078 | $2.09 \times 10^{-4}$ | Data not available | | No instruments | | No instruments | | no |
| AGER<br>4125-52_2 | -0.030 | 0.141 | -0.189 | $2.70 \times 10^{-2}$ | No instruments | | -0.041 | $6.09 \times 10^{-5}$ | yes |
| DPT<br>4979-34_2 | 0.019 | 0.367 | 0.109 | 0.192 | -1.519 | 0.114 | -0.023 | $1.24 \times 10^{-4}$ | yes |
| CTSA<br>3179-51_2 | 0.036 | $8.40 \times 10^{-2}$ | 0.209 | $1.70 \times 10^{-2}$ | 0.672 | 0.320 | -0.075 | $1.18 \times 10^{-7}$ | yes |

In the protein‐to‐BMI direction, we found that the 2SMR was more powered than the 1SMR. This is also plausible and could be attributed to the fact that individual‐level genetic associations with BMI as an outcome are much weaker in a study the size of KORA, while the effect estimates from larger GWAS with BMI [2, 3] are much more precise. In all applicable cases (including nominal associations), we found consistency in the MR effect directions between the 1SMR and 2SMR, and in both directions of the MR (Supplementary Table 9, 10, 11, 12). We therefore focus our analysis on 1SMR in the BMI‐to‐protein direction and on 2SMR in the protein‐to‐BMI direction.

### BMI is potentially causal for 24 of the 152 tested proteins

The 1SMR approach in the BMI‐to‐protein direction allowed investigation of potentially causal relationships between BMI and 152 replicated blood plasma proteins. BMI was suggested to have a causal effect on 24 proteins, after correction for multiple testing (Figure 5, Supplementary Table 9).

**Figure 5:**
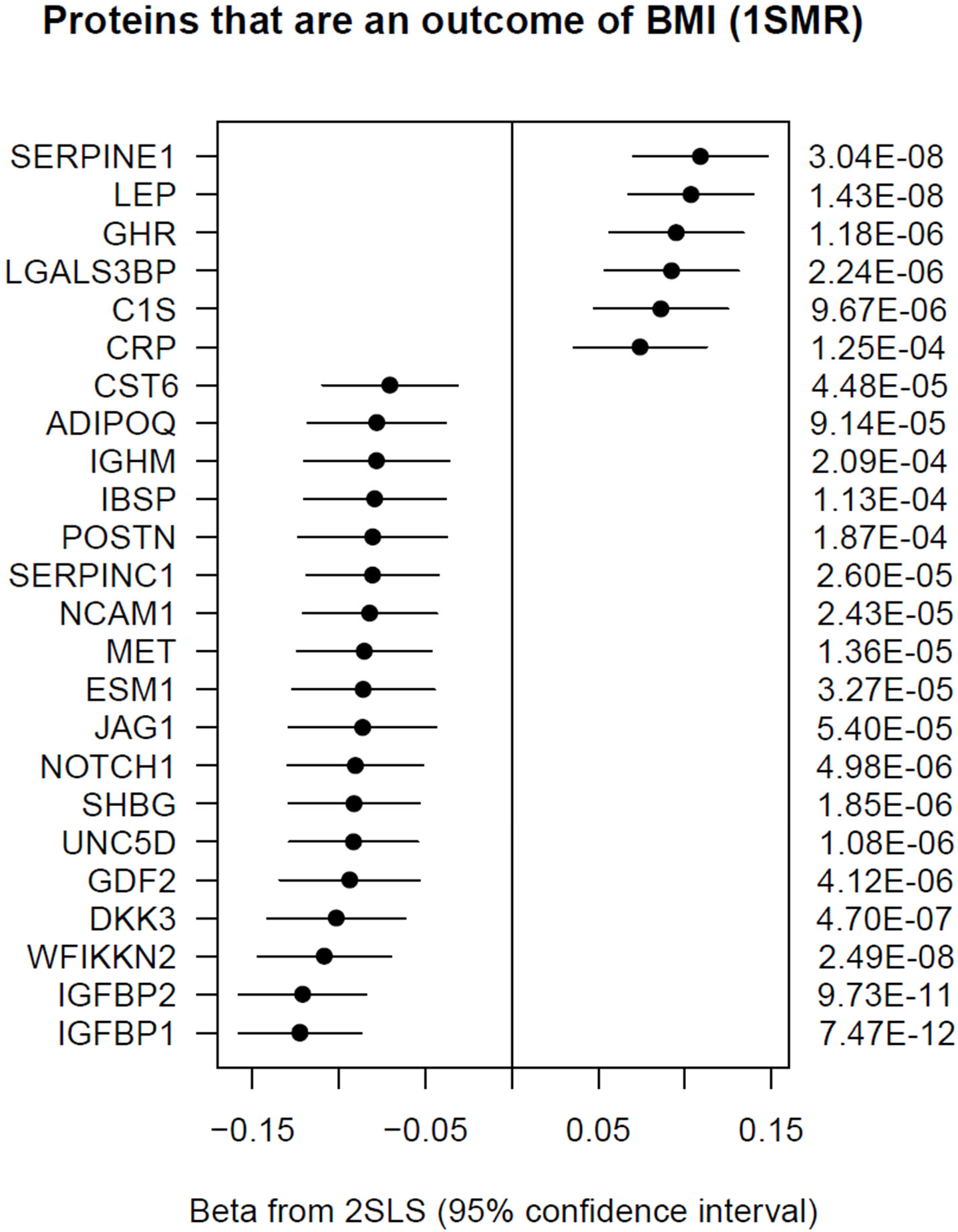
Forest plot of the causal estimate of BMI on various proteins in the one-sample MR analysis (KORA). BMI is suggested to have a causal effect on 24 out of 152 replicated proteins, using the 2SLS method. The BMI polygenic score (GPS_BMI_) was used as an instrument for BMI in this analysis.

### Six plasma proteins have a potentially causal role in the development of obesity

Using 2SMR analysis in the protein‐to‐BMI direction, we used the Proteome PheWAS browser [18] which curated SNPs associated with proteins from five protein GWASs [13, 16, 19‐21] and categorized protein instruments based on their suitability for MR analysis. We identified genetic instruments for 82 of the 152 replicated proteins, in addition to leptin receptor (LEPR), which we considered a positive control in this study. This analysis suggested that six proteins (LEPR, IGFBP1, WFIKKN2, AGER, DPT, and CTSA) may potentially have a causal role in the development of obesity, after correction for multiple testing (p<0.05/82 = 6.10×10^‐4^) (Supplementary Table 12).

In summary, we found that BMI had a causal effect on the levels of 24 proteins, while six proteins had a potentially causal role in the development of obesity, three of which are suggested to have a role in both directions (LEPR, IGFBP1, and WFIKKN2).

### Biological role of the 28 causally and/or consequentially BMI‐associated proteins

To study the tissue‐specific role of the causal/consequential proteins in obesity, we screened the Genotype‐Tissue Expression (GTEx) human database and the Mouse Genome Informatics (MGI) database. We found that the 28 proteins can be clustered into two groups (Supplementary Figure 4a). The first cluster showed wide‐spread expression across 54 different human tissues and similarly across various mouse tissues. For instance, NCAM1, DKK3, IGFBP2, LGALS3BP, SERPINE1, and NOTCH1 were predominantly expressed in brain, adipose and heart tissues. The second cluster showed more sporadic expression in relevant human tissues, such as ADIPOQ and LEP in adipose tissue, and IGFBP1, CRP and SERPINC1 in liver tissue, CST6 is expressed in skin, and WFIKKN2 in ovaries, testis, and brain. In mice, WFIKKN2 is primarily expressed in the brain, heart, eyes, and pancreas (Supplementary Figure 4b). There is not much knowledge about the role of WFIKKN2 in obesity, however it is known to have a regulatory role of some members of the transforming growth factor beta (TGFB) family [22]. The TGFB superfamily are produced in adipose tissues and involved in the regulation of adiposity [23] and obesity is known to alter their expression level.

We extensively searched the literature for animal models for the 28 proteins (Supplementary Table 13). We found biological evidence for 18 protein coding genes, out of 28 linked with obesity, in animal models. Some of the causal proteins, in particular, AGER knock out models showed accelerated weight gain and increased plasma cholesterol [24], and CTSA knock out models presented in having thick skin that contained enlarged hyperplastic epidermal glands as well as a reduction in dermal fat [25]. The circulating soluble receptor for advanced glycation end products (AGER) is negatively associated with BMI [26], as we also observed in KORA (beta = ‐1.09 and p = 2.42×10^−13^). In addition, recent evidence suggests a novel role of the adipokine dermatopontin (DPT) in obesity by regulation of adipose tissue remodeling and inflammation [27]. A DPT knockout mouse model showed increased subcutaneous adipose tissue [28], and others showed effects on skin elasticity, dermis thickness, and collagen accumulation.

We further investigated the 28 proteins for enrichments in obesity traits, using the hybrid mouse diversity panel (HMDP) [29] and secondly using an F2 cross of the inbred ApoE-/- C57BL/6J and C3H/HeJ strains [30] (see Methods). 26 of the proteins had mouse orthologs [31]. We observed correlations (R>0.1 and p-value<0.05) between adipose and liver tissue gene expression and numerous essential obesity traits, including body fat composition, bone density, insulin, and various lipid traits such as LDL, HDL, cholesterol, triglycerides, etc. (Figure 6).

**Figure 6:**
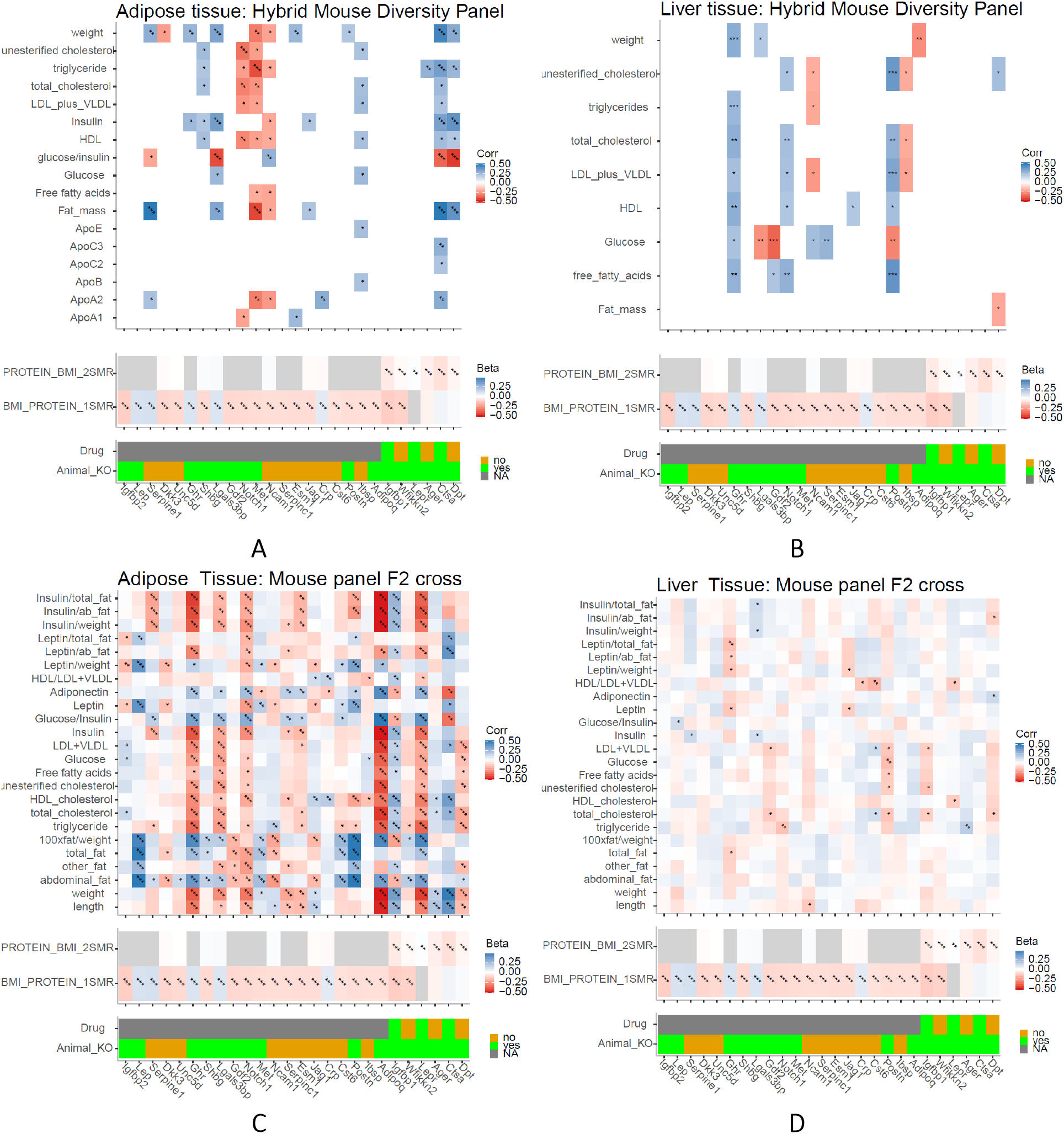
Adipose and liver tissue gene expression associations with obesity traits in mouse panels. The bi-weight mid-correlation coefficients (median-based measures of similarity) and p-values are shown for obesity-related traits with adipose/liver tissue gene expression levels using a threshold of p<0.05 and absolute correlation coefficient > 0.1 in two datasets: (A,B) the HMDP dataset consisting of 706 mice fed a standard chow diet and (C,D) the F2 dataset which is a cross of the inbred ApoE-/- C57BL/6J and C3H/HeJ strains fed a high fat + cholesterol diet. The significance of the correlations is as indicated (*** for p<0.001, ** for p<0.01, and * for p<0.05).

Lastly, we used the DrugBank database [32] to search for existing drugs that target the six proteins that were causal for BMI. Three proteins were targets for at least one existing drug that has completed phase II clinical trials (Supplementary Table 14). This included Metreleptin [33] which is a target for leptin receptor to treat complications of leptin deficiency in individuals with congenital or acquired lipodystrophy. Another drug, Pegvisomant [34], is a highly selective growth hormone receptor antagonist that is used to treat acromegaly by production of IGF-1 which is the main mediator of growth hormone activity. A third drug, Mecasermin [35], targets IGFBP1 and IGFBP2 by acting as an agonist of insulin-like growth factor 1 receptor. It is a drug that is used for the treatment of growth failure in pediatric patients with primary IGFD or GH gene deletion.

## Discussion

### Proteins associated with obesity and obesity score

We identified 152 proteins that were significantly associated with BMI in KORA and confirmed in at least one other study. We then applied a systematic approach that was derived and validated in a previous study [17] to compute a GPS_BMI_ in approximately 1,000 individuals from the KORA study. The genetic background of the KORA participants is similar to the cohort on which the score computation was based (both European) [2]. The GPS_BMI_ was strongly associated with BMI and also accurately showed differences in leptin levels, a protein whose association with BMI and obesity is well established, among numerous other obesity-related proteins.

This is the first study to test a genetic score that was based on European ancestry in a population of Arabs and mixed ethnicities. This may have yielded a weaker association with BMI in QMDiab compared to the European cohort (p = 5.54×10^−4^), but the statistical power of QMDiab is also more limited by the cohort size (N = 353). It is therefore difficult to interpret whether generalization of the GPS_BMI_ score was limited due to differences in the genetic architecture of obesity between the populations or due to the sample size.

The 19 proteins that were associated with GPS_BMI_ displayed an amplified effect for the individuals in the tail of the GPS_BMI_ distribution. These proteins all have important roles in obesity that have been well documented in the literature. Proteins of the insulin-like growth factor system and their receptors including insulin-like growth factor-binding protein 1 and 2 (IGFBP1 and IGFBP2), and growth hormone (GH) have central pathophysiological roles in normal metabolism, insulin resistance, and type 2 diabetes [36], mainly by modulating insulin sensitivity. IGFBP2 has been shown to be regulated by leptin [37]. Growth hormone also has an important role in the development of obesity and GH receptor (GHR) mutations were reported in obese individuals [38]. Netrin receptor (UNC5D), is another mediator of inflammation known to promote macrophage retention in adipose tissue [39]. Markers of high adiposity also include increased concentrations of CRP [40] [41] and SERPINE1 [42], contrasted by reduced levels of SERPINC1 [43] and SHBG [44], all of which we observe in this study.

### Causal analysis

Our study suggests that BMI has a potentially causal effect on 24 proteins. This is in line with a previous study that reports widespread effects of adiposity on DNA methylation [7]. On the other hand, our 2SMR also suggests that LEPR, IGFBP1, WFIKKN2, AGER, DPT, and CTSA are potentially causal candidates for obesity. Taken together, our data suggests that a bi-directional relationship is likely and may be reciprocated in other BMI-protein associations due to the underlying complexity of the disease and multitude of involved pathways.

### Animal models

Mice studies showed that LEPR knockout mice became excessively obese [45]. LEP and LEPR deficient mice have also been shown to be hyper-insulinemic, hyperglycemic (depending on the age and strain), and having elevated total cholesterol levels with a predominance of LDL/HDL1 particles [46–48]. LEP and LEPR levels may be both a sensor of fat mass and at the same time, part of a negative feedback mechanism to maintain a set point for body fat stores [49]. In some instances, such as leptin deficiency in monogenic obesity, the causal direction in one direction is obvious [50]. Further suggesting the causal role proteins may have on BMI, a global IGFBP1 deletion in mice showed a significant increase in body weight and body fat mass [51]. Epigenetic regulation of IGFBP2 in abdominal obesity [52] also points towards having a causal role.

### Other biological evidence

WAP, Kazal, immunoglobulin, Kunitz and NTR domain-containing protein 2 (WFIKKN2) is a protease-inhibitor that contains multiple distinct protease inhibitor domains [53]. WFIKKN2 encodes GASP1 (Growth and differentiation factor-associated serum protein-1) [54] and is an inhibitory TGF-β binding protein. It has mainly been involved in skeletal and muscle fiber development in the heart [55]. Higher WFIKKN2 protein levels were associated with lower levels of fasting insulin, triglycerides, HOMA-IR and visceral fat [56] suggesting a protective role against metabolic dysregulation.

In addition, global overexpression of GASP1 resulted in a significant increase in body weight in mice [57]. Interestingly, our findings were consistent with a recent SomaLogic protein study of type 2 diabetes in AGES- Reykjavik, where WFIKKN2 was reported to be potentially causal for type 2 diabetes [58], independent from BMI. Furthermore, WFIKKN2 was suggested as a potentially causal candidate for type 2 diabetes, in a second study from INTERVAL that associates the diabetes risk score with proteins [59], after adjusting for age, sex and technical covariates. Lastly, genetic variants in the WFIKKN2 locus (cis-pQTLs) showed regulation of GDF8/11 at the protein level in a trans-pQTL manner [16]. Thus, the plasma levels of GDF8/11 and WFIKKN2 are strongly controlled by genetics. Genetically supported targets could be twice as successful as those without genetic support in clinical practice [60], suggesting that WFIKKN2 is a potential target that would modulate GDF8/11 function.

## Conclusion

Genome-wide polygenic scores capture genetic susceptibility by aggregating effects of genome-wide variation with individually modest effects. The cumulative genetic effects captured in these scores regulate the plasma proteome and can influence the development of obesity. We mined our GPS to protein associations to determine which protein levels would associate with genetic predisposition to obesity. Then, by modeling polygenic scores as proxies for obesity, we have identified putatively causal effects of BMI on 24 plasma proteins using MR. In the reverse direction, we identified six plasma proteins with a causal effect on BMI. Complementing our MR findings with existing animal model data, our data suggests that LEPR, WFIKKN2, and IGFBP1 are both a readout and driving factor for obesity, while AGER, DPT, and CTSA have a predominantly causal effect on BMI. Thus, our computational approaches combined with assimilated experimental data, coherently suggest that the revealed associations can be bi-directional rather than strictly uni-directional. By overlaying our causal proteins with both human and mouse gene expression information and experimental evidence, we highlight potential new drug targets for follow-up studies.

## Methods

### Study population (KORA)

The KORA F4 study is a population-based cohort of 3,080 subjects living in southern Germany. Study participants were recruited between 2006 and 2008 comprising individuals with age ranging from 32 to 81. Other covariates that were considered included binary diabetes information (case/control based on self reporting or medication usage), physical activity, alcohol consumption, and smoking. For this study aptamer-based proteomics was done using the SOMAscan platform the protein levels of 996 individuals and has been described in detail elsewhere [13].

### Proteomics (KORA)

The SOMAscan platform was used to quantify protein levels of 996 KORA individuals. Details of the SOMAscan platform have been described elsewhere [61–66]. Briefly, undepleted EDTA-plasma is diluted into three dilution bins (0.05, 1, and 40 %) and incubated with bin-specific collections of bead-coupled SOMAmers in a 96-well plate format. Subsequent to washing steps, bead-bound proteins are biotinylated and complexes comprising biotinylated target proteins and fluorescence-labelled SOMAmers are photo cleaved off the bead support and pooled. Following recapture on streptavidin beads and further washing steps, SOMAmers are eluted and quantified as a proxy to protein concentration by hybridization to custom arrays of SOMAmer-complementary oligonucleotides. Based on standard samples included on each plate, the resulting raw intensities are processed using a data analysis work flow including hybridization normalization, median signal normalization and signal calibration to control for inter-plate differences. One-thousand blood samples from the KORA F4 study were sent to SomaLogic Inc. (Boulder Colorado, USA) for analysis. Of the original 1,000 samples three did not have BMI information and one sample failed SOMAscan QC, leaving a total of 996 samples. Data for 1,129 SOMAmer probes (SOMAscan assay V3.2) was obtained for these samples. 29 of the probes failed SOMAscan QC, leaving a total of 1,100 probes for analysis.

### Genotyping *(KORA)*

The Affymetrix Axiom Array was used to genotype 3,788 samples of the KORA S4 of which 996 were used in this study. After thorough quality control (total genotyping rate in the remaining SNPs was 99.8%) and filtering for minor allele frequency (MAF) > 1%, a total of 509,946 autosomal SNPs was used for imputation. Shapeit v2 was used to infer haplotypes from the available SNPs using the 1000G phase 3 haplotype build 37 genetic maps. Impute2 v2.3.2 was used for imputation. Variants with certainty < 0.95, information metric<0.7, missing genotype data (geno 0.2), Hardy-Weinberg equilibrium (hbe) exact test p-value < 1×10^−6^, or with MAF <0.01 were all excluded. A total of 8,263,604 variants with a total genotyping rate of 0.97 were kept for the analysis.

### Study population (QMDiab)

The Qatar Metabolomics Study on Diabetes (QMDiab) is a cross-sectional case-control study that was carried out in 2012 at the Dermatology Department in Hamad Medical Corporation (HMC Doha, Qatar). This cohort was described previously and comprises 388 study participants from Arab and Asian ethnicities of which around 50% have type 2 diabetes [15]. A subset of 356 samples having proteomics data were used in this study.

### Proteomics (QMDiab)

The SOMAscan platform of the WCM-Q proteomics core was used to quantify a total of 1,129 protein measurements in 356 plasma samples from QMDiab [13]. Protocols and instrumentation were provided and certified using reference samples by SomaLogic Inc. No samples data or probes were excluded.

### Genotyping (QMDiab)

Genotyping was carried out using the Infinium Human Omni 2.5-8 V1.2 Beadchip array for 353 samples and was previously described elsewhere [13]. After stringent quality control, 1,221,345 variants were used to impute a total of 18,829,416 variants that were used in this study. The same imputation quality metrics were used in QMDiab as in KORA.

### Polygenic score calculation

Polygenic scores represent a quantification of an individuals inherited risk by combining the impact of thousands of common variants. Derivation, validation, and testing of the score is described elsewhere [17] [2] [67]. Briefly, the score was derived using summary statistics from a recent GWAS study for BMI covering up to 339,224 individuals and a reference panel of 503 European samples from 1000 Genomes phase 3 version 5 Ambiguous SNPs (A/T or C/G) were not included in the score derivation. A set of candidate scores were derived using the LDPred algorithm [68] which is a Bayesian method and pruning and threshold derivation strategies. Another approach that involved pruning and thresholding was used to derive additional candidate scores using an LD-driven clumping procedure in PLINK version 1.90b (--clump). These scores were then validated in another dataset. The scores were generated by multiplying the dosage of each risk allele for each variant by its respective weight, and summing across all variants in the score, while incorporating genotype dosages for the uncertainty in genotype imputation. Finally, the optimal score having the best discriminative capacity based on highest AUC with BMI as the outcome in the UK Biobank validation dataset was selected.

The derived weights of the optimal candidate BMI score were used to generate BMI scores for the 996 samples from KORA and the 353 samples from QMDiab. Scoring was carried out using PLINK version 2.0 [69]. Imputed genotyping data was used, and in total, 1,583,718 and 1,636,172 variants passed QC for the GPS computation, in KORA and QMDiab respectively. Finally, the GPS_BMI_ values were scaled to have a mean of 0 and standard deviation of 1.

### Statistical Analysis

The protein measures were log2 transformed and standardized (mean = 0, sd = 1) in both KORA and QMDiab. For the BMI-protein associations in KORA, linear models were used while adjusting for age and sex. Another linear model that adjusts for age, sex, smoking, alcohol, physical activity, and diabetes was also evaluated in KORA as a sensitivity analysis. For replication of the BMI-protein associations in QMDiab, analysis was performed using a slightly different model: (age + sex + study-specific covariates). The study-specific covariates in QMDiab consisted of diabetes status, the first three principal components (PCs) of the genotyping data (genoPC1, genoPC2, and genoPC3) along with the first three principal components of the proteomics data (somaPC1, somaPC2, and somaPC3) which were added as covariates in the analysis. Diabetes was used as a covariate in QMDiab to eliminate associations confounded by the diabetes-BMI relationship. These PCs were considered as standard covariates of the QMDiab study [15]. The genetic PCs accounted for the ethnic variability of the QMDiab cohort and the proteomics PCs accounted for a moderate level of observed cell lysis. Finally, the association of proteins with the BMI scores was carried out also using linear regression while adjusting for age and sex in KORA, and adjusting for age, sex, genetic PCs, and soma PCs in QMDiab.

To consider a BMI-protein association as “replicated,” we required p<2.72×10^−4^ (0.05/184). We also estimated the statistical power for the replication by sampling. This was carried out for each association by randomly selecting 356 individuals from the KORA cohort (without replacement) and computing the p-value of association for that subset of samples. This was repeated 1,000 times and 50^th^ smallest p-value from this distribution was considered to be obtained at 95% power (p95).

## Mendelian Randomization analysis

Causal inference was carried out using 1SMR and 2SMR. The majority of MR analyses were conducted using the “MendelianRandomization” v0.4.1 [70] or the “TwoSampleMR” v0.4.22 [71].

To assess the effect of BMI on proteins in the 1SMR, we modeled the GPS_BMI_ as an instrument for BMI using KORA data. We used the 2SLS method to compute the causal estimate of BMI on to the 152 replicated proteins, while adjusting for age and sex. This was performed using the “ivreg” function from the “ivpack” R package v.1.2.

To assess the effect of BMI on protein levels, in a 2SMR setting, we identified BMI instruments in the GIANT-UK Biobank meta-analysis [3], and extracted the corresponding SNP-exposure estimates from the INTERVAL pGWAS [16]. We selected instruments for BMI to have genome-wide significance (p<1×10^−8^ and f-statistic>10) and an LD clumping threshold of 0.001. 454 BMI instruments were used after these filtration steps and further eliminating SNPs with potential confounders from the UK Biobank GWAS [72]. The exposure and outcome data were harmonized before performing the MR analysis by aligning the SNPs on the same effect allele for the exposure (BMI) and outcome (proteins). The 2SMR was feasible for 103 of the 152 replicated proteins for which GWAS summary statistics were available from INTERVAL. We used the IVW method to estimate the causal effect of BMI on proteins. We downloaded full protein GWAS summary statistics from INTERVAL, and extracted the genetic instrument SNPs as outcome associations from this data. The 2SMR results using the IVW method, suggested that BMI was causal for nine of the 103 tested proteins, after multiple-testing correction. The causal estimates were directionally concordant with the 1SMR estimates for all significant proteins. These results were also robust to sensitivity analysis and there was also no strong evidence of heterogeneity or horizontal pleiotropy based on the MR Egger analysis (Supplementary Table 10).

For the 1SMR in the protein-to-BMI direction, we identified protein instruments in KORA and also computed the corresponding SNP-outcome exposure estimates for the 152 proteins. For each protein, instruments were tested for association with protein levels in a linear regression model adjusted for age and sex. Genetic instruments were selected if their association was genome-wide significant (f-statistic>10). The genome-wide instruments were filtered to include only independent signals (r2 = 0.001). Out of the 152 plasma proteins, we identified one or two suitable genetic instruments for 63 proteins. After accounting for multiple testing, the 1SMR did not provide any evidence of plasma proteins having a potentially causal effect on BMI, due to lack of strong/suitable instruments (Supplementary Table 11).

Finally, for the 2SMR in the protein-to-BMI direction, the Proteome PheWAS browser http://www.epigraphdb.org/pqtl/ accessed on April 2020) [18] was used to check if any of the proteins with suitable instruments were causal for BMI. Instrument reliability was based on pleiotropy, consistency, and colocalization scores, as defined by the authors of that study. With a single genetic variant, the estimate of the IVW reduces to the ratio of coefficients betaY/betaX or the Wald ratio.

## Gene expression analysis

To investigate the tissue-specific role of the GPS_BMI_ associated protein coding genes with obesity, we used RNA-seq tissue expression data from both human and mice - the Genotype-Tissue Exxpression (GTEx) database [73] and the Gene eXpression Database (GXD) [74] (C57BL/6J strain), respectively. The data presented and described in this manuscript were generated through a multi-gene query on the GTEx portal on 03/29/2020 from: https://www.gtexportal.org/home/multiGeneQueryPage. Mice expression data was visualized on the GXD portal where expression data was processed using the Morpheus heat map and visualization and analysis tool created by the Broad Institute from http://www.informatics.jax.org/expression.shtml.

## Identifying the tissue-specific role of GPS-associated proteins in obesity using mouse databases

Mouse orthologs were identified for the genes encoding the causal/consequential proteins using the Mouse Genome Informatics (MGI) database (http://www.informatics.jax.org/) [31]. An ortholog was present for 26 proteins. Two reference mouse databases were used to identify correlations between adipose and liver tissue expression of these proteins and the relevant obesity traits in mice: a) the hybrid mouse diversity panel (HDMP; n = 706 mice from 100 well-characterized inbred strains fed with a standard chow diet) [29], and b) an F2 cross of the inbred ApoE-/- C57BL/6J and C3H/HeJ strains (n = 334 mice that were fed with a high fat and cholesterol diet from 8–16 weeks of age and sacrificed at 24 weeks of age) [30]. Publically available data from the systems genetic resource was downloaded and used to search for gene-trait correlations in adipose and liver tissues from https://systems.genetics.ucla.edu/[29]. The adipose and liver tissue expression data of the 334 F2 cross mice was accessed using the publically available dataset Sage BioNetworks at https://www.synapse.org/#!Synapse:syn4497 [30]. The biweight midcorrelation coefficients (bicor) is a similarity measure between samples based on median which is less sensitive to outliers and provides a robust alternative to similarity metrics like Pearson correlation. The bicor coefficients and the p-values for the association of gene expression levels and the selected obesity relevant traits were computed using the WGCNA R package [75]. Gene to trait correlations were filtered to only include absolute correlation coefficients>0.1 and p-value<0.05 for both datasets.

## Data Availability

All summary statistics and association data for KORA and QMDiab are available in Supplementary Tables 2, 3, 5, and 8. The informed consent given by the study participants does not cover posting of participant level phenotype and genotype data in public databases. However, data are available upon request from KORA-gen (http://epi.helmholtz-muenchen.de/kora-gen). Requests are submitted online and are subject to approval by the KORA board.

## Acknowledgements

The KORA-Study Group consists of A. Peters (speaker), J.Heinrich, R.Holle, R.Leidl, C.Meisinger, K.Strauch and their co-workers, who are responsible for the design and conduct of the KORA studies. We gratefully acknowledge the contribution of all members of field staff conducting the KORA F4 study. We thank the staff of the HMC dermatology department and of WCM-Q for their contribution to QMDiab. The Genotype-Tissue Expression (GTEx) project was supported by the Common Fund of the Office of the Director of the National Institutes of Health, and by NCI, NHGRI, NHLBI, NIDA, NIMH, and NINDS. Finally, we are grateful to all study participants of KORA and QMDiab for their invaluable contributions to this study.

## Ethics approval and consent to participate

Project agreement for this study was granted under K060/18g. All KORA participants have given written informed consent and the study was approved by the Ethics Committee of the Bavarian Medical Association. The QMDiab study was approved by the Institutional Review Boards of HMC and WCM-Q under research protocol number 11131/11).

All study participants provided written informed consent.

## Author contributions

Conceived and designed the study: SBZ, SS, HG, KS

Performed experiments: SBZ, SS

Analyzed data: SBZ, SS, HG, KS

Contributed reagents/materials/analysis tools: MM, PRM, MAE, WR, JG, KS, AP, CG, MW

Wrote the paper: SBZ, SS, HG, KS

All authors discussed the results and reviewed the final manuscript.

## SUPPLEMENTARY FIGURES

**Supplementary Figure 1:**
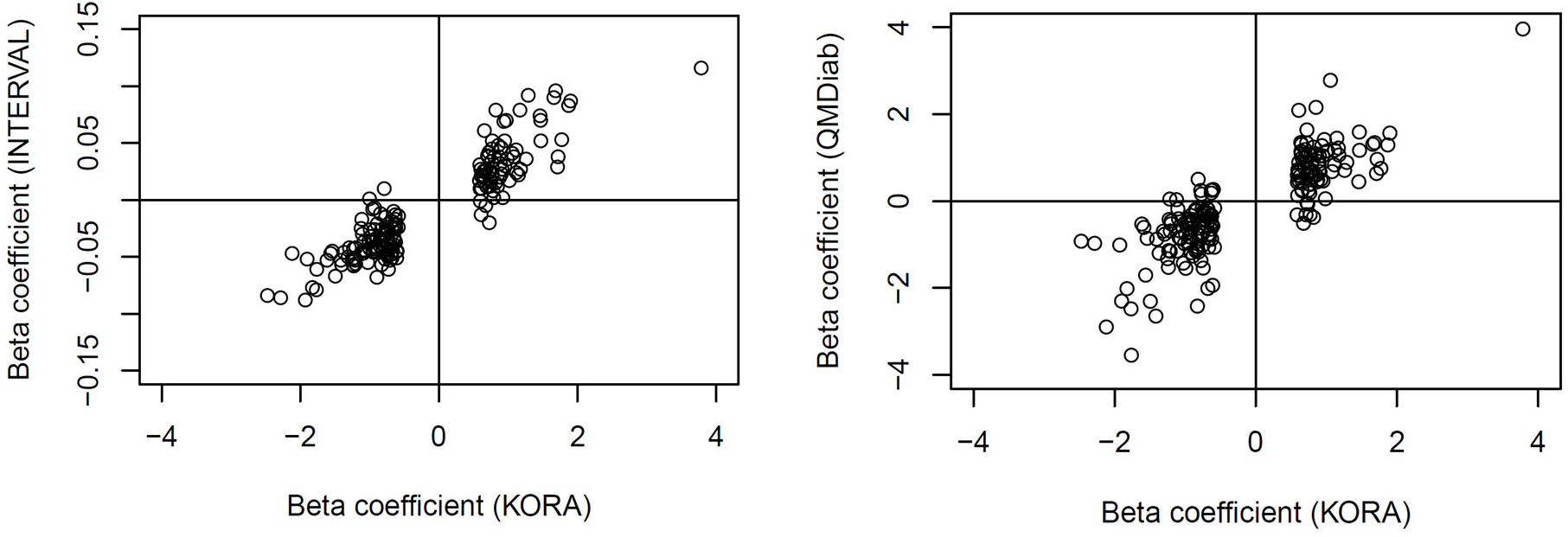
Scatterplot of the regression coefficients from the three studies. The effect sizes of the BMI-protein associations for the 184 proteins that are significant in KORA are compared to the observed effects in INTERVAL and QMDiab.

**Supplementary Figure 2:**
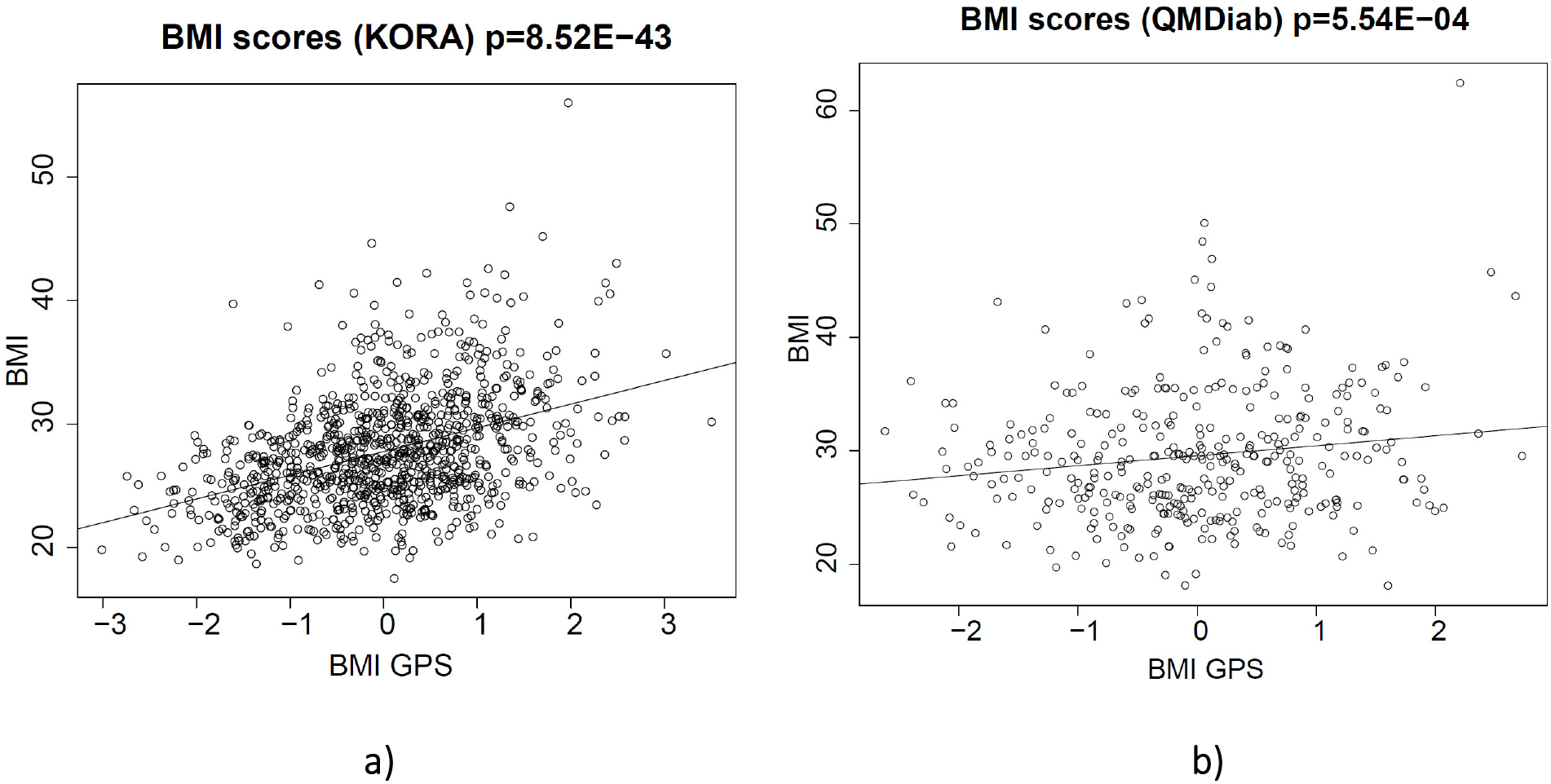
The computed GPS_BMI_ strongly associates with BMI. a) KORA and b) QMDiab.

**Supplementary Figure 3:** Tail-effect for GPS_BMI_ and 18 blood circulating proteins (3a-3r). Different effect sizes and significance levels at various percentiles of the GPS_BMI_ distribution were compared and showed an over-proportionally increased genetic predisposition for developing obesity in the extreme tail.

**Supplementary Figure 4:**
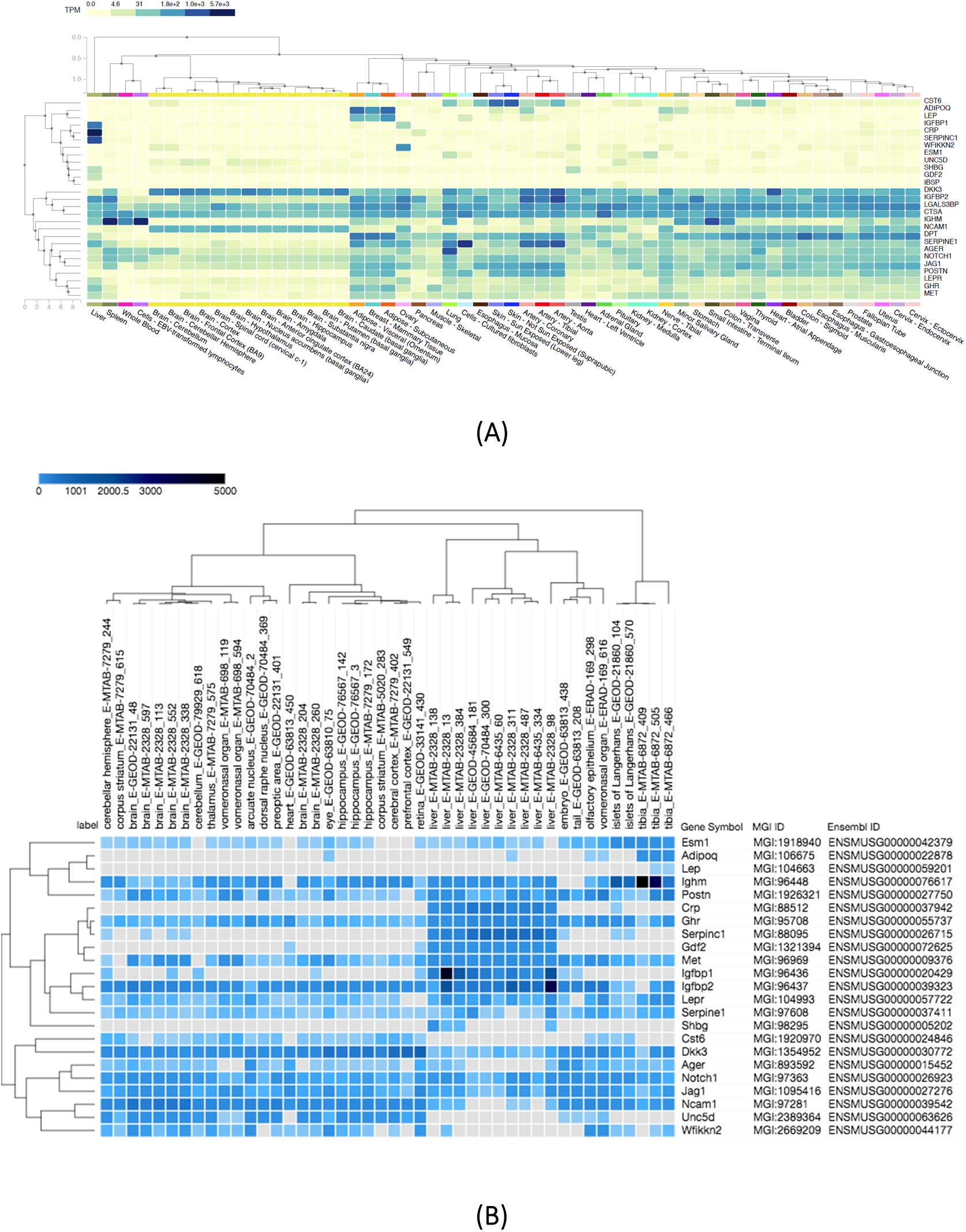
Human and mice tissue-specific gene expression and regulation for the causal/consequential proteins. A) Human RNA-seq data from GTEx showing the transcript per million (TPM) expression values for the genes encoding the proteins. Data is arranged into two clusters. B) RNA-Seq tissue specific gene expression and regulation in mice for the C57BL/6J strain from the MGI database. The expression values for the genes encoding the proteins are shown in transcript per million (TPM). Data is only shown for the available genes in the respective databases.

**ST1: KORA descriptive statistics**. We provide the study descriptive statistics for the 996 individuals included in this study. We tested whether there were any significant differences (p<0.001) in any of the available phenotypes (sex, age, alcohol consumption, smoking status, physical activity, and blood biomarkers) between the obese (BMI>= 30) and the overweight (BMI<30) individuals.

**ST2: Association of BMI with proteins (simple model).** Association statistics for the association of BMI with proteins in a linear regression model lm(BMI ∼ proteins + age + sex). Columns O-AI include the SOMAscan assay v4 annotations version 3.3.2, Column AJ includes information on cross-reactive proteins from Sun et al. (INTERVAL pGWAS study), where a subset of the Somalogic aptamers (SOMAmers) were tested for cross-reactivity with homologous proteins that have at least 40% sequence similarity. Columns AK-AQ contain assessment of the specificity of the SOMAlogic assay for the given proteins provided from the Emilsson et al. AGES pGWAS study. Direct assessment of aptamer specificity was carried out using data dependent analysis (DDA) and multiple reaction monitoring (MRM) mass spectrometry after SOMAmer enrichment in biological matrices.

**ST3: Association of BMI with proteins (full model).** Association statistics for the association of BMI with proteins in a linear regression model lm(BMI ∼ proteins + age + sex + alcohol + physical activity + smoking + diabetes).

**ST4: Replication of KORA results in both INTERVAL and QMDiab.** Columns A-D contain the protein information. Columns E-G contain the association statistics for the association of BMI with proteins in a linear regression model lm(BMI ∼ proteins + age + sex) in KORA. Columns H-J and K-M contain the same statistics in INTERVAL and QMDiab respectively. Columns N-Q include information on concordance of directionality and significance level of the replication.

**ST5: Association of BMI GPS with proteins in KORA.** Association statistics for the association of BMI GPS with proteins in a linear regression model lm(BMI_GPS ∼ proteins + age + sex).

**ST6: Replication of BMI GPS - protein associations in QMDiab.** Association statistics for the association of BMI GPS with proteins in a linear regression model with QMDiab specific paramters: lm(BMI_GPS ∼ proteins + age + sex + genoPCs + somaPCs).

**ST7: Table comparing GPS-protein statistics when the GPS computation includes/excludes cis-SNPs.** There is no considerable difference in significance p-value when including cis-SNPs in the BMI score computation.

**ST8: Association of proteins with different GPS extremes of population.** Association statistics for the association of BMI GPS with proteins in a linear regression model lm(BMI_GPS ∼ proteins + age + sex) for different GPS extremes.

**ST9: One-sample MR using UKBB-based polygenic BMI score as instrument in KORA BMI-->Protein (2SLS).** The 2SLS method was used to obtain the causal estimate of BMI on the 152 replicated proteins (associated with BMI), while adjusting for age and sex. 24 plasma proteins were Bonferroni significant after correction for multiple testing p<0.05/152.

**ST10: Two-sample MR (BMI-->Protein) using inverse variance weighted method.** Here we show the MR statistics used to determine if BMI has a causal effect on the observed protein levels. Instruments for BMI were selected to have genome-wide significance (p<1E-8 and f-statistic>10) and an LD clumping threshold of 0.001. Full protein GWAS summary statistics for 103 of the 152 replicated proteins, were available from the Sun et al. INTERVAL pGWAS, from which the genetic instrument SNPs were selected as outcome associations. To assess the causal estimate of BMI on the 103 proteins, for which data was available, we used the inverse-variance weighted method (IVW). The results of the two-sample MR using the IVW method, suggest that BMI has a causal effect on nine of the tested proteins, after multiple-testing correction (Bonferroni p<0.05/103) (indicated in red color). BMI shows a causal effect for an additional 10 proteins that were significant in the 1SMR, at nominal signficance (p<0.05). On the right side, we compare the IVW method (purple) to the MR Egger method (green). The estimate from the Egger method is consistent with the IVW estimate for all proteins shown below. The Cochran’s Q statistic (I.sq.egger) represents the heterogeneity (a measure of the degree to which genetic instruments identify the same causal effect). The significance of this heterogeneity is represented by p_Het and a low p-value corresponds to evidence of non-directional pleiotropy. P_pleio is the p-value for the MR-Egger intercept test and a low p-value corresponds to evidence of directional pleiotropy.

**ST11: One-sample MR using in KORA Protein --> BMI (2SLS)**. The 2SLS method was used to obtain the causal estimate of the 152 replicated proteins (associated with BMI) on BMI, while adjusting for age and sex. Out of the 152 plasma proteins, we identified suitable genetic instruments for 63 proteins, none of which the MR p-value was significant. However, we show the MR statistics for the proteins that were significant in any of the other MR analyses for comparison purposes.

**ST12: Two-sample MR (Protein-->BMI) using Proteome PheWAS browser.** To study the direction (protein levels being causal to BMI) in a two-sample MR analysis, we used the Proteome PheWAS browser http://www.epigraphdb.org/pqtl; accessed on April 2020) which curated SNPs associated with proteins from five protein GWASs. Genetic instruments with information on their quality metrics were identified in the PheWAS browser for 82 of the 152 replicated proteins, in addition to leptin receptor which was not included in our protein panel. SNP reliability was determined based on the pleiotropy score, consistency, and colocalization test, Tier 1 being the most and Tier 3 being the least reliable. Tier 1 instruments passed both pleiotropy and consistency tests, and were considered primary instruments for the MR analysis. Tier 2 instruments showed evidence of high heterogeneity across studies either the pairwise X test (Z>5) or colocalization analysis (PP<80%). Tier 3 instruments were those associated with more than 5 proteins and are considered non-specific (highly pleiotropic) and were excluded from all analyses. The results of this two-sample MR analysis suggest that these six proteins may potentially be causal for BMI, after Bonferroni correction for multiple testing (p<0.05/82). Note: With a single genetic variant, the estimate of the IVW reduces to the ratio of coefficients betaY/betaX

**ST13: Animal models for causal/consequential proteins.**

**ST14: Drug target information.** Data is extracted from the DrugBank database (https://www.drugbank.ca accessed on April 2020) on existing drugs that target any of the six causal proteins for obesity.

